# Treatment Outline and Clinical Outcome of Hospitalized COVID-19 Patients: Experiences from a Combined Military Hospital of Bangladesh

**DOI:** 10.1101/2022.03.01.22271740

**Authors:** Sabiha Mahboob, Fatema Johora, Asma Akter Abbasy, FatihaTasmin Jeenia, Mohammad Ali, Md Humayun Kabir, Ferdaush Ahmed Sojib, Jannatul Ferdoush

## Abstract

**Background:** Global knowledge of treatment and outcomes of COVID-19 has been evolving since the onset of the pandemic.

**Materials and Methods:** The objective of this cross-sectional study was to explore treatment and outcome of COVID-19 patients admitted in a Combined Military Hospital of Bangladesh. Data were collected from treatment records of patients of the CMH Bogura during the period of June 2020 to August 2020. Total 219 RT-PCR positive admitted patients were included as study population.

**Result:** Among 219 patients, 78.6% were male and 21.5% were female, mean age of patients was 34.3 ± 12.2. About14.6% patients had one or more comorbidities. Most (83.1%) of the admitted patients were diagnosed as mild cases. Antimicrobials were used in 98.8% cases, and frequent use of doxycycline (80.4%) and ivermectine (77.2%) was found. Anticoagulant and steroid therapy were used in 42.0% and 15.5% patients respectively. O_2_ therapy was required in 6.0% cases and intensive care unit (ICU) support was needed in 2.3% cases.Duration of hospital stay was 12.1± 4.4 days and 100% of patients were discharged from hospital. There was no single mortality during the study period.

**Conclusion:** High prevalence of antimicrobials use was observed among the hospitalized COVID-19 patients in this single center study.Supportive care was effective with no incidence of mortality.

## INTRODUCTION

Coronavirus disease 2019 (COVID-19), the highly contagious viral illness caused by severe acute respiratory syndrome coronavirus 2 (SARS-CoV-2), has had a catastrophic effect worldwide.After the detection of the first case of this predominantly respiratory viral illness in Wuhan, Hubei Province, China, in late December 2019, SARS-CoV-2 rapidly disseminated across the world in a short span of time.^1, 2^ The World Health Organization (WHO) has declared it as a global pandemic on March 11, 2020. Since being declared a global pandemic, the COVID-19 pandemic emerged as a major public health emergency affecting the healthcare services all over the world.^3, 4^

Coronavirus disease 2019 (COVID-19) is characterized by hyperinflammatory response, vascular damage, microangiopathy, angiogenesis and widespread thrombosis.^5^ Four stages of COVID-19 have been identified. The first stage is characterized by upper respiratory tract infection, the second by the onset of dyspnea and pneumonia, the third by a worsening clinical scenario dominated by a cytokine storm and the consequent hyperinflammatory state, and the fourth by death or recovery.^6^The complex pathophysiology, pulmonary and extrapulmonary disease, and immune mediated effects make medical management more challenging than many viral illnesses.^7^Following infection with SARS-CoV-2, some diseased individuals may remain asymptomatic or only present with mild upper respiratory symptoms, or may develop pneumonia and severe acute respiratory distress syndrome (ARDS) that requires intubation in intensive care and presents complications with an ominous outcome.^5,8^

Currently, no treatment is very effective for prevention and treatment of SARS-CoV-2 infection. Several vaccines are available all over the world. Vaccination reduces the chance of infection as well as mortality and morbidity.^9,10^ The COVID-19 nationwide immunization campaigns in Bangladesh began on February 7, 2021, based on a set of criteria that prioritized the most vulnerable members of the population.^11^ Based on the pathological features and different clinical stages of COVID-19, the classes of drugs that are mainly used include steroids, low-molecular-weight heparins antiviral agents, plasma, and hyperimmune immunoglobulins.^5,7,12,13^ Unlike influenza infections and related pneumonia, a few COVID-19 patients experience an initial bacterial superinfection at admission due to pneumococci, other streptococci or staphylococci and require antimicrobial therapy.^14,15^Management and outcome of COVID-19 varied from country to country.^16^There are few available data in this contextfrom Bangladesh.^17,18,19^Hence, the present study was conducted to discuss treatment outline and clinical outcome ofCOVID-19 patientsadmitted in a combined military hospital of Bangladesh.

## MATERIALS &METHODS

This retrospective cross-sectional study was conducted in a Combined Military Hospital (CMH) Bogura of Bangladesh from June 2020 to August 2020 with the aim to explore treatments and outcomes of admitted COVID-19 patients. Ethical approval was taken from Institutional Review Board (IRB) of CMH Bogura. Data were collected from treatment records of COVID-19 patients of CMH Bogura during the period of June 2020 to August 2020. Investigators collected retrospective data from chronological register and treatment records kept in the Record Room of hospital. Data collection started from the most recently registered in-patient records to gradually backward manner. RT-PCR positive admitted COVID-19 patients of all age groups and sexes were included as study population. RT-PCR negative but radiologically suggestive cases of COVID-19 patients were excluded from the study. Treatment sheets of total 219 patients were included in this study. A review form was used for data collection. For maintenance of anonymity, an identification number was provided at top of the front page of the treatment sheet where confirmed clinical diagnosis and patient profile were mentioned. Data was compiled, presented and analyzed using Microsoft Excel 2007, and was expressed as percentage.

## Results

**Table I** showing out of 219 patients, 78.6% were male and 21.5% were female.Mean age (years) of patients was 34.3 ± 12.2.Most (85.4%) of the patients had no comorbidity.

**Table I.** Demographic characteristics of admitted COVID-19 patients.

| Gender | Frequency | Percentage |
| --- | --- | --- |
| Male | 172 | 78.6 |
| Female | 47 | 21.5 |
| Age (years) | Mean ± SD | |
| | $34.26 \pm 12.13$ | |
| Comorbidities | Frequency | Percentage |
| Present | 32 | 14.6 |

Most (83.1%) of the hospitalized patients were diagnosed as mild cases (**Figure 1**).

**Figure1:**
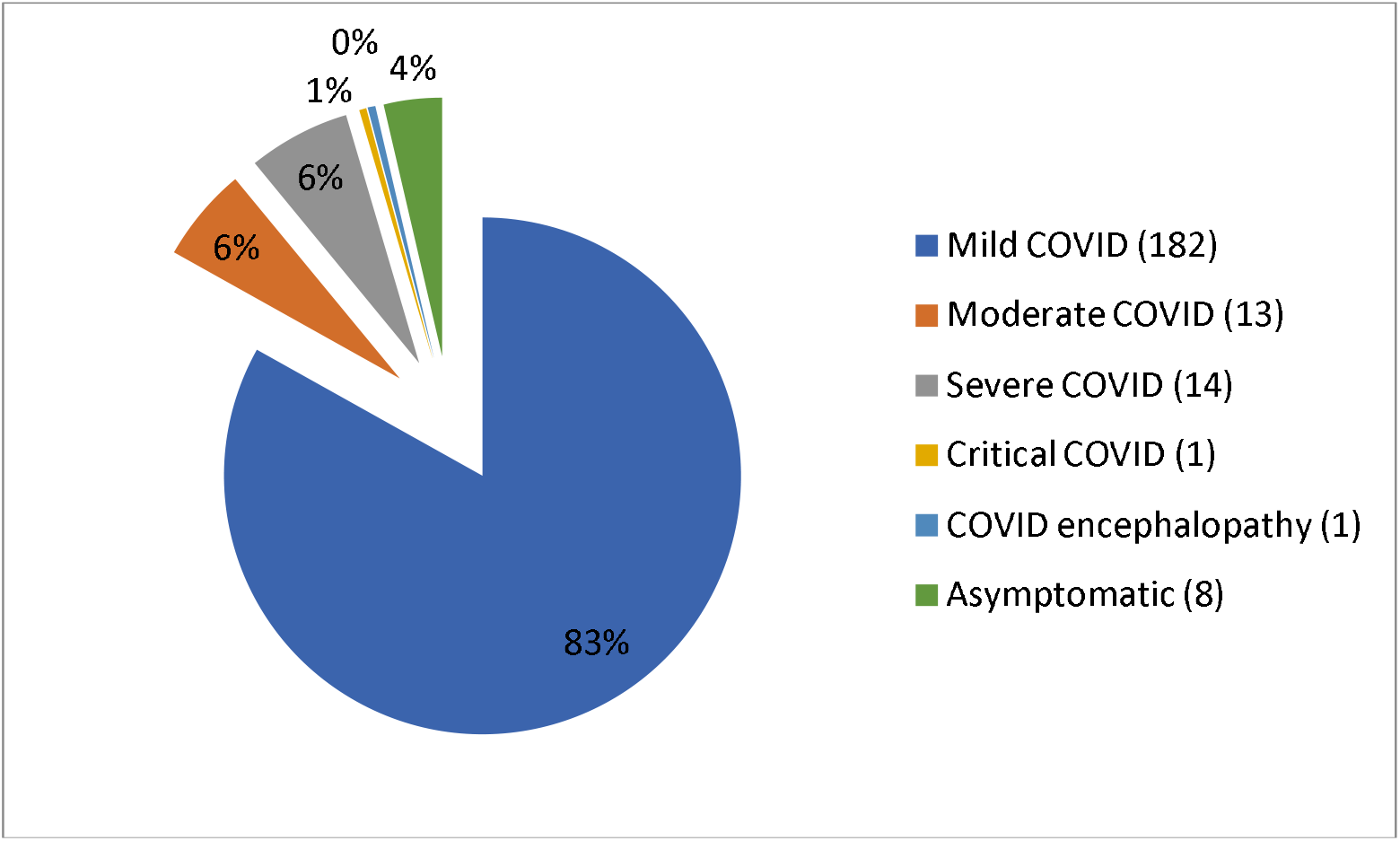
Diagnosis of hospitalized COVID-19 patients.

**Table II.**
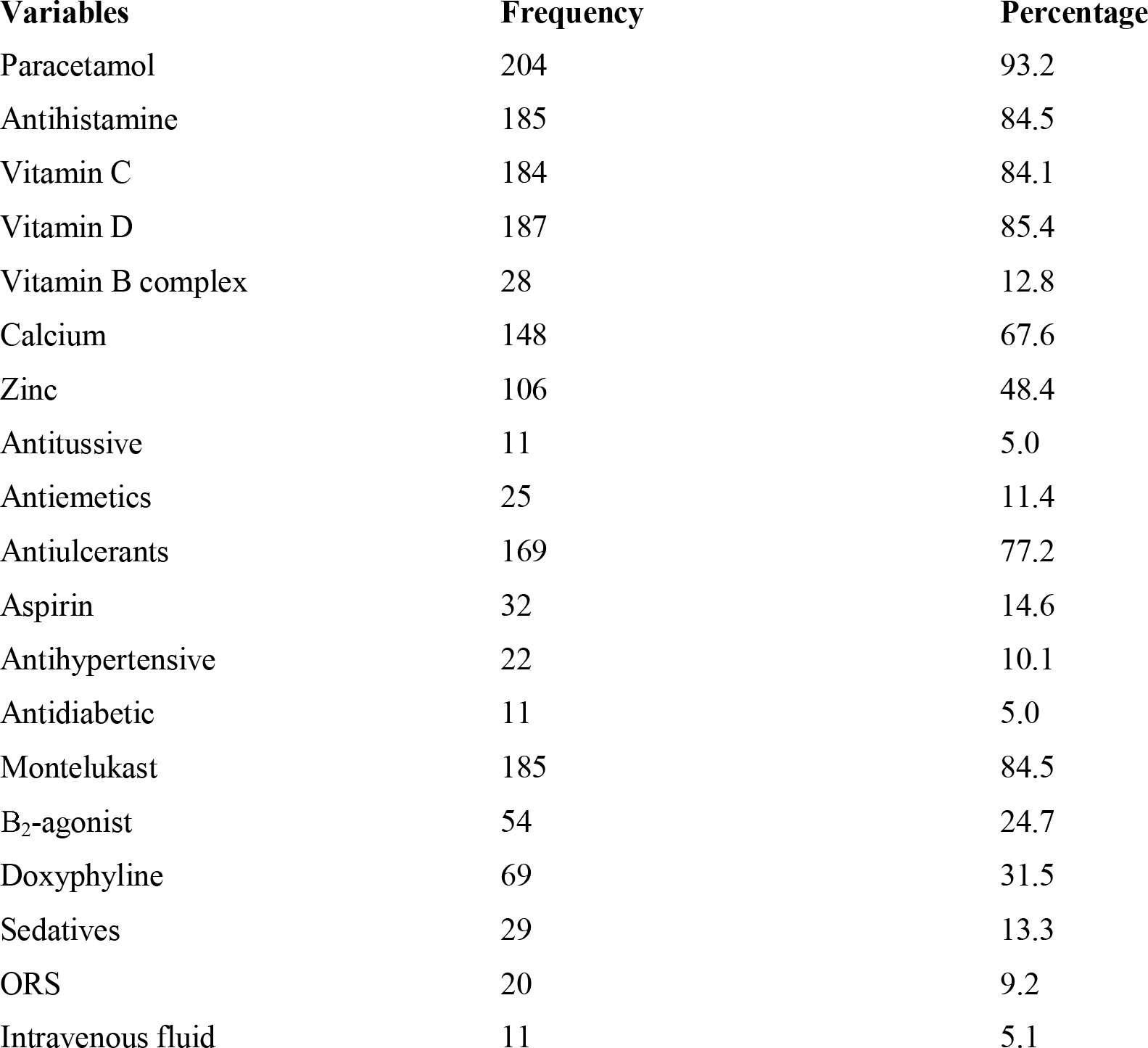
Treatment details of COVID-19 patients.

Most (98.8%) of the patients received 1 or more antimicrobial agents. 42.0% patients received anticoagulant drugs, where 15.5% received steroid therapy.

**Figure 3** showed that doxycycline (80.4%) was the frequently used antimicrobials followed by ivermectine (77.2%). Other antibiotics (amoxillin, ceftriaxone, cefuroxime, meropenem, levofloxacin, moxifloxacin etc) were used in alone or combination in 35.6% cases.

**Figure 2:**
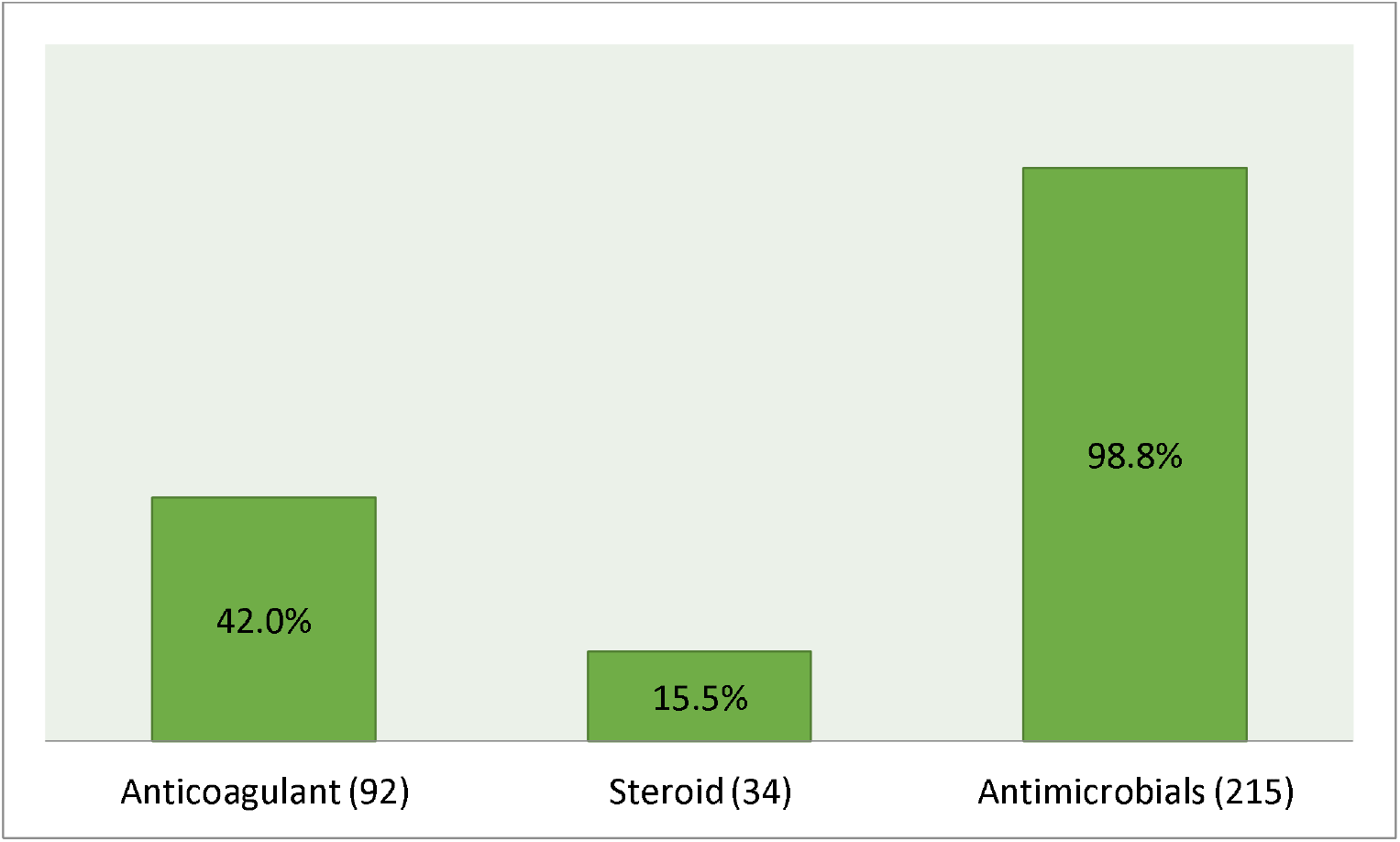
Proportion of patients received anticoagulant, steroids and antimicrobial agents.

**Figure 3.**
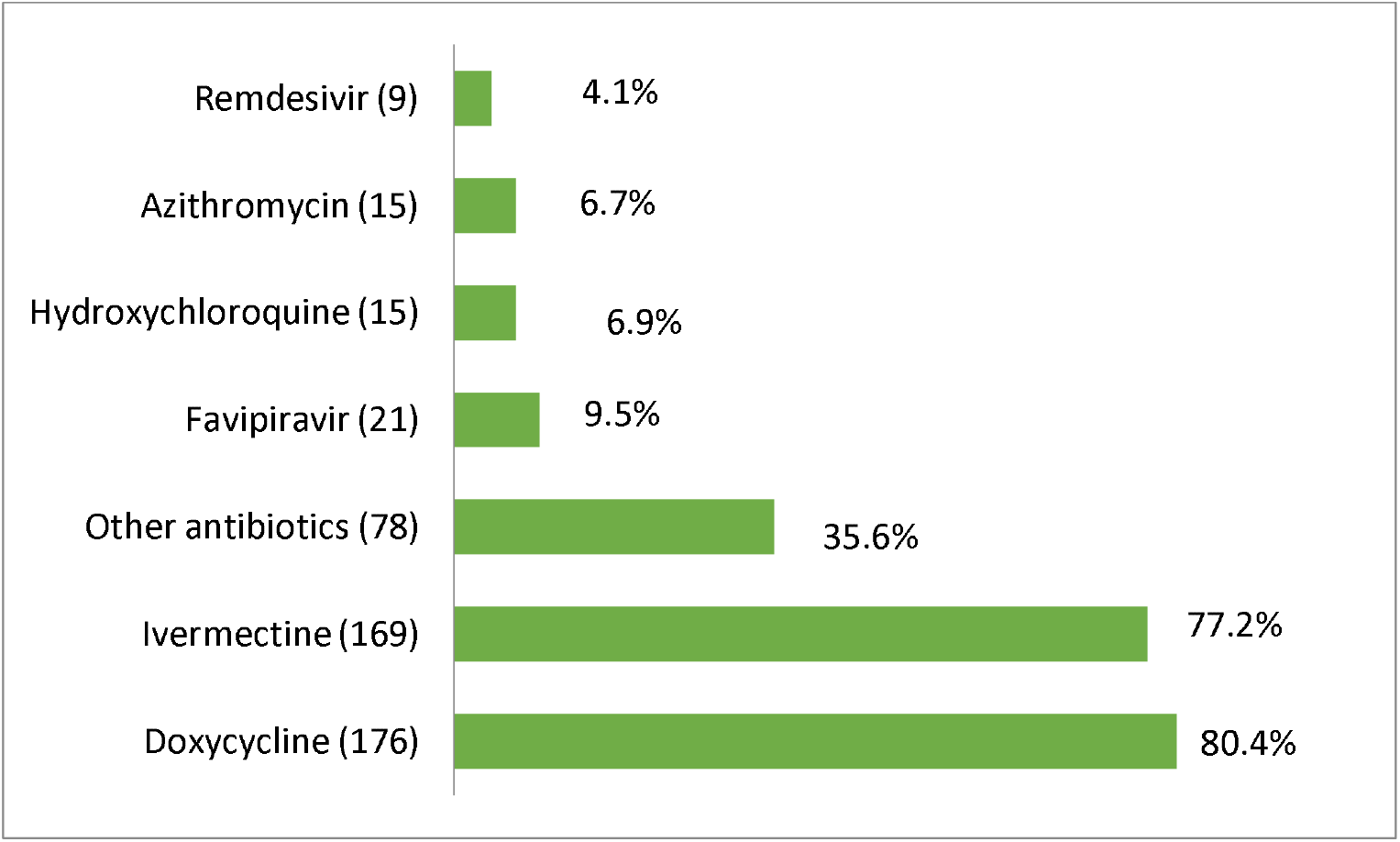
Proportion of antimicrobials used in COVID-19.

Only 6.0% of patients required O_2_ therapy and 2.3% patients were needed intensive care unit (ICU) support **(Figure 4 & 5)**.

**Figure 4.**
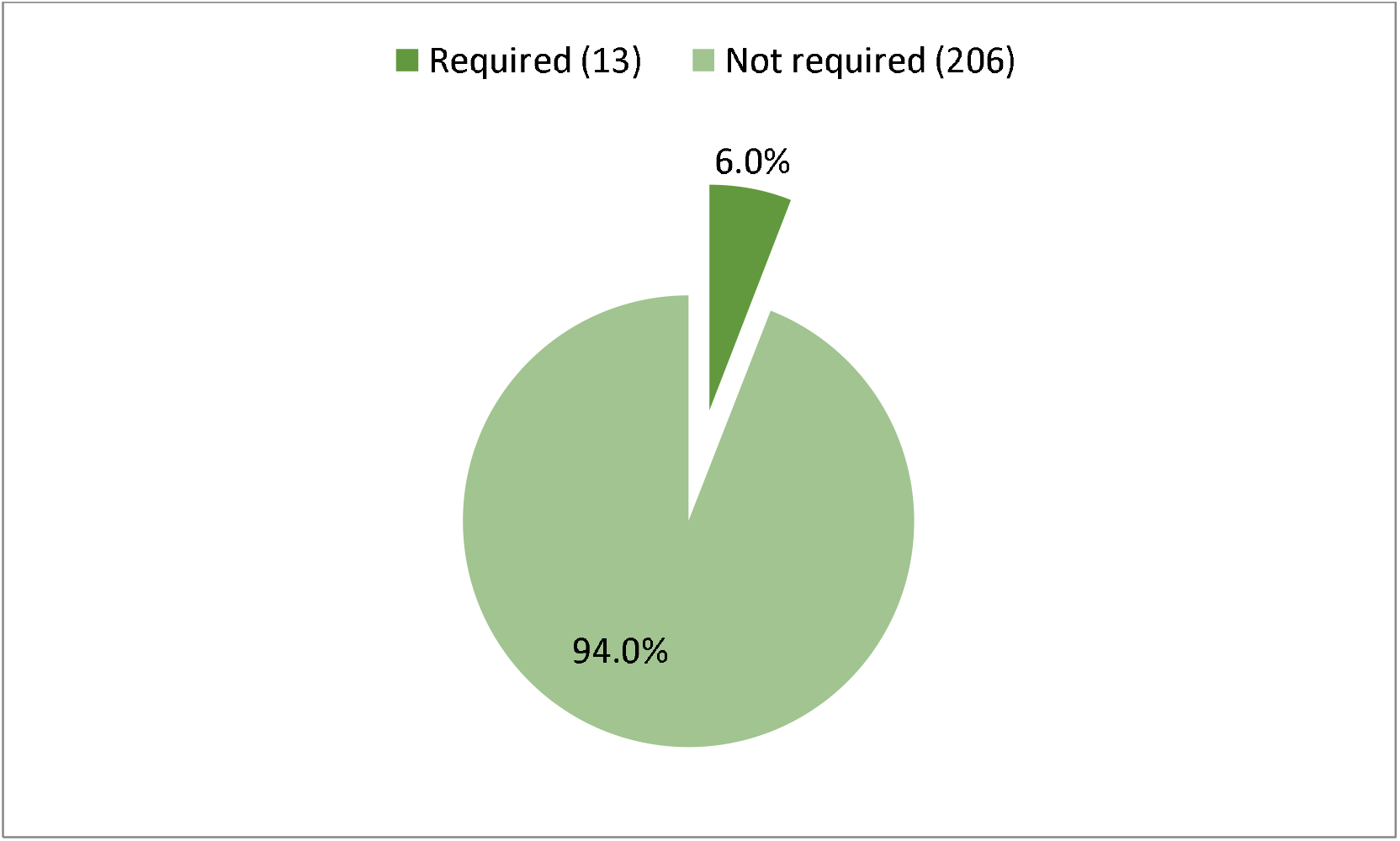
Proportion of patients required O_2_ therapy.

**Figure 5.**
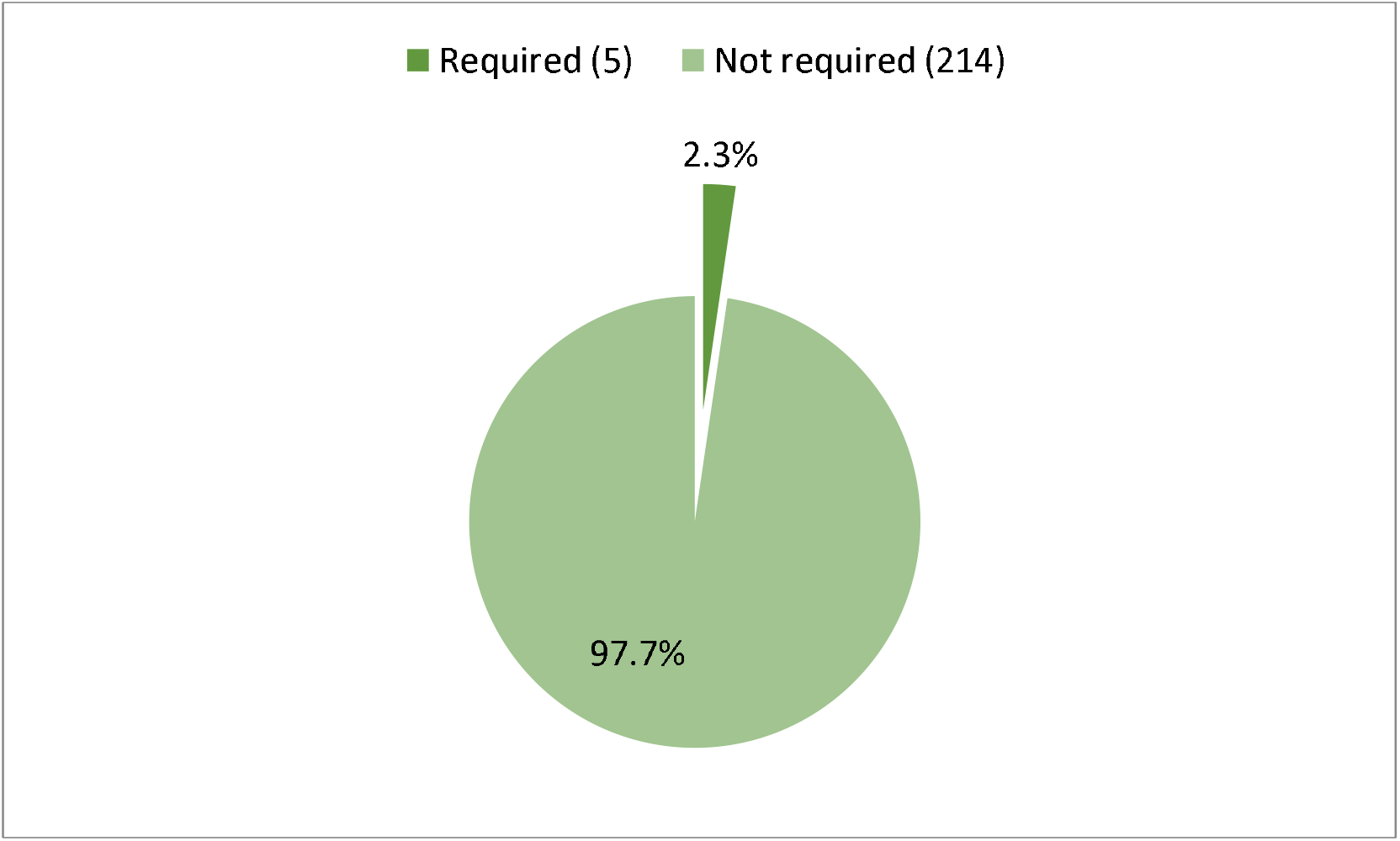
Proportion of patients required intensive care unit (ICU) support.

**Table III** showed duration of hospital stay was 12.1± 4.4 days. All 219 patients (100%) were discharged from hospital. There was not a single case of death or referring to higher center during the study period.

**Table III.** Hospital course and outcome of admitted COVID-19 patients.

| **Variables** | **Mean ± SD** |  |
| --- | --- | --- |
| **Duration of hospital stay (days)** | 12.1± 4.4 | |
| **Outcome** | **Frequency** | **Percentage** |
| Discharge | 219 | 100 |
| Transferred out | 0 | 0 |
| Death | 0 | 0 |

## DISCUSSION

For several months, COVID-19 pandemic has put enormous burden on healthcare system all over the world. Highly contagious nature, complex pathogenicity and absence of a definitive therapeutic agent all are making this pandemic a long run for human being.^20^Initially physicians tried ‘repurposing’ of several existing therapeutic molecules along with symptomatic treatment to combat COVID-19. At the same time, several clinical trials were conducted to study the safety and effectiveness of several repurposed drugs for COVID-19 treatment.^21^ Later on countries were started to develop clinical guidelines for management of coronavirus with the support of World Health Organization.^13^ Current study was conducted to assess treatment outline and clinical outcome of hospitalized COVID-19 patients of a combined military hospital during the first wave of COVID-19 pandemic.

Present study revealed that 98.8% of the admitted patients received one or more antimicrobial agents although most of the cases were mild disease. Despite of no recommendation of antimicrobial for treatment of mild cases in clinical guidelines, higher usage of antimicrobials in COVID-19 patients was observed in previous studies done in home and abroad.^13,18,22,23,24^Increased prescribing of antimicrobials in this pandemic is alarming.^25^COVID-19 patients could receive antimicrobial therapy for several reasons. Firstly, clinical scenario of COVID-19 is confusing and difficult to distinguish from bacterial pneumonia. As a result, almost 70% of hospitalized COVID-19 patients receive empiric broad-spectrum antibiotic therapy due to lack of confirm microbiological diagnosis. Secondly, prevention of presumed secondary bacterial infections is another reason of antimicrobials prescribing in COVID-19 patients although 16% of hospitalized develop secondary bacterial infection, where antibiotics are needed. Injudicious use of antimicrobials effective against MRSA or antipseudomonal antimicrobials in criticalCOVID-19 patients could worsen the current situation of AMR.^26^ Doxycycline and ivermectine were frequently used antimicrobials and this finding is contrary to related literatures as previous researchers found azithromycin as frequently prescribed agent.^24,27^ Ahmed et al described the administration of either hydroxychloroquine, doxycycline, azithromycin or favipiravir alone or in combination to all hospitalized patients in another combined military hospital of Bangladesh.^19^ One study from United States found significantly increased dispensing of ivermectin although it was not is not authorized or approved by FDA for prevention or treatment of COVID-19.^28^ Nasir et al revealed ivermectin and doxycycline were frequently consumed as self-medication during the first wave of pandemic. Lower usage of remdesivir, favirapir and hydroxychloroquine was observed in current study that was similar to previous studies done in different countries including Bangladesh.^22,24,27,30,31^But two recentstudies done in Bangladesh found higher usage of these agents in certain percentage of patients having comorbidities.^17,18^

Steroids have the potential to prevent an extended cytokine response and may accelerate resolution of pulmonary and systemic inflammation in pneumonia. Methylprednisolone, may improve dysregulated immune response caused by sepsis (possible complication of infection with COVID-19) and hypotension. Dexamethasone has been reported to reduce the duration of mechanical ventilation. Long-term glucocorticoid therapy has displayed significant improvement in indices of alveolar–capillary membrane permeability and mediators of inflammation and tissue repair, and reduction of duration of mechanical ventilation. In this study, steroid was prescribed to moderate, severe or critical COVID patients and this practice was concordance with national guideline.^13^ Two similar studies done in Bangladesh revealed higher usage of steroid. Increased age, comorbidities and severity of disease were seen among the population of those two studies.^17, 18^

Based on available evidence, it is reasonable to administer venous thromboembolism prophylaxis with either a low molecular weight or unfractionated heparin in hospitalized patients. In patients with rapidly progressing respiratory deterioration or where clinical judgment suggests thrombosis, treatment doses of anticoagulants may be considered.^32,33^In this study,42% patients received anticoagulant therapy and that was similar to Soni et al.^34^ Where study done by Islam et al showed 100% patients had thromboprophylaxis as they treated severe and critical illness more than this studied hospital.^17^

Montelukast, a leukotrine receptor antagonist, mainly used as a complementary therapy in adults in addition to inhaled corticosteroids in bronchial asthma and COPD.^35^It is also known to decrease the frequency and severity of wheezing after an upper respiratory tract infection caused by adenovirus, influenza, metapneumovirus or coronavirus.^36^More effectiveness could be find in patients with comorbidities such as bronchial asthma, diabetes, sleep apnea, smoking, obesity, or symptomatic atherosclerotic lesions.^35^ In current study, 84.5% patients received montelukast although only 14.6% had comorbidities. There was no available study to compare this finding.There was no recommendation of montelukast in national guideline.^13^ Antipyretics, antihistamines as symptomatic treatment and calcium, zinc, vitamin C, vitamin D were used as supplemental therapy in this study like other related studies. ^17, 18, 19, 34^

Present study showed that 5.9% patients required oxygen therapy and that was similar to one study conducted in Bangladesh.^19^Higher requirement of oxygen was seen in other studies conducted in Bangladesh and India.^18,34,39^In this study, 2.3% patients required intensive care and that was concurrent to three relevant studies. ^19, 34, 37^ But findings of Ali et al (8.7%), Soni et al (52%) and Hossain et al (24.9%) were different from current study.^18,34,39^Younger age group, less comorbidityand mild casesmight be the reason of less requirement of oxygen therapy and intensive care in our study.^19,36^

In current study, duration of hospital stay was 12.1 ± 4.4 days and that was different from earlier study conducted in Bangladesh.Islam et al showed hospital stay was less than 10 days in maximum cases although most of the cases were severe and critical.^17^Despite having 83% mild cases with longer hospital stay indicates difference in policy of studied hospitals. In our study, there was no single incidence of mortality, 100% patients were discharged from hospital. 0.03% and 1% mortality were observed in two studies conducted in Bangladesh.^18,19^Another two studies done in Bangladesh found 4.7% and 19.3% mortality respectively.^17,39^1.4% and 2.6% mortality were observed in Indian studies.^34,37^Younger age group, less comorbidity and mild form of disease favor good outcome of our patients.^17,39^

## CONCLUSION

High prevalence of antimicrobials usewas observedamong the hospitalized COVID-19 patients in current study.Supportive care was effective with no incidence of mortality.

## Data Availability

All data produced in the present study are available upon reasonable request to authors

## REFERENCE

1. Zhu, N, Zhang D, Wang W, Li, et al. A Novel Coronavirus from Patients with Pneumonia in China, 2019. The New England Journal of Medicine2020; 382(8):727–33

2. Zhou P, Yang XL, Wang XG, et al. A pneumonia outbreak associated with a new coronavirus of probable bat origin. Nature 2020;579(7798): 270–3.

3. World Health Organization (WHO). WHO Director-General’s opening remarks at the media briefing on COVID19 -March 2020. WHO, Geneva, Switzerland. Available at: https://www.who.int/director-general/speeches/detail/who-director-general-s-opening-remarks-at-the-press-conference---march-2020 [Accessed on 10.02.2022]

4. Sharma A, Tiwari S, Deb MK, et al. Severe acute respiratory syndrome coronavirus-2 (SARS-CoV-2): a global pandemic and treatment strategies. International Journal of Antimicrobial Agents 2020; 56 (2):106054.

5. Basher A, Das D, Faiz M.A. Coronavirus Disease 2019 (COVID-19) – An Overview. J Bangladesh Coll Phys Surg2020; 38: 84–90

6. Stasi C, Fallani S, Voller F, et al. Treatment for COVID-19: An Overview. European Journal of Pharmacology 2020; 889:.173644–173644

7. Matricardi PM, Dal Negro RW, Nisini R. The first, holistic immunological model of COVID-19: Implications for prevention, diagnosis, and public health measures. Pediatr Allergy Immunol. 2020;31(5):454–470.

8. World Health Organization (WHO). Clinical management of COVID-19. World Health Organization, Geneva, Switzerland. Available on: http://WHO-2019-nCoV-clinical-2020.5-eng.pdf [Accessed on 10.02.2022]

9. World Health Organization (WHO). Immunization in the context of COVID-19 pandemic. World Health Organization, Geneva, Switzerland. 2020. Available at: https://www.who.int/publications/i/item/immunization-in-the-context-of-covid-19-pandemic. [Accessed on 10.02.2022]

10. World Health Organization (WHO). Accelerating a safe and effective COVID-19 vaccine. World Health Organization, Geneva, Switzerland.2020. Available at: https://www.who.int/emergencies/diseases/novel-coronavirus-2019/global-research-on-novel-coronavirus-2019-ncov/accelerating-a-safe-and-effective-covid-19-vaccine [Accessedon 10.02.2022]

11. World Health Organization (WHO). COVID-19 vaccines. World Health Organization, Geneva, Switzerland. 2021.Available at: https://www.who.int/emergencies/diseases/novel-coronavirus-2019/covid-19-vaccine [Accessedon 10.02.2022]

12. McFee RB. COVID-19: Therapeutics and interventions currently under consideration. Dis Mon. 2020; 66(9):101058. doi:10.1016/j.disamonth.2020.101058

13. Directorate General of Health Services (DGHS). National Guidelines on Clinical Management of Coronavirus Disease 2019 (COVID-19). DGHS, Ministry of Health & Family Welfare, Dhaka, Bangladesh. 2020. Available at: https://mofa.portal.gov.bd/sites/default/files/files/mofa.portal.gov.bd/page/ad1f289c_47cf_4f6c_8dee_887957be3176/National%20Guidelines%20on%20Clinical%20Management%20of%20Covid-19-%20DGHS.pdf[Accessed on 10.02.2022]

14. Rawson TM, Moore LSP, Zhu N,, et al. Bacterial and fungal co-infection inindividuals with coronavirus: A rapid review to supportCOVID-19 antimicrobial prescribing. Clin Infect Dis.2020; 71(9):2459–2468

15. Westblade LF, Simon MS, Satlin MJ. Bacterial coinfections in coronavirus disease 2019. Trends Microbiol. 2021;29(10):930–941.

16. Jamison DT, Lau LJ, Wu KB, et al. Country performance against COVID-19: rankings for 35 countries. BMJ Global Health 2020; 5:e003047.

17. Islam QT, Hossain H, Fahim F, et al. Clinico-Demograhic Profile, Treatment Outline and Clinical Outcome of 236 Confirmed Hospitalized COVID-19 Patients: A Multi-Centered Descriptive Study in Dhaka, Bangladesh. Bangladesh Journal of Medicine, 2020; 31(2), 52–57.

18. Ali MR, Hasan MA, Rahman MS, et al. Clinical manifestations and socio-demographic status of COVID-19 patients during the second-wave of pandemic: A Bangladeshi experience. J Infect Public Health. 2021;14(10):1367–1374.

19. Ahmed NU, Islam MA, Kabir MA, et al. Clinico-Pathological Findings of Bangladeshi Covid 19 Patients with their Clinical Outcome: Study of A Cohort of 201 Cases. Journal of Bangladesh College of Physicians and Surgeons 2020; 38, 37–42.

20. Muralidar S, Ambi SV, Sekaran S, et al. The emergence of COVID-19 as a global pandemic: Understanding the epidemiology, immune response and potential therapeutic targets of SARS-CoV-2. Biochimie 2020; 179:85-100.

21. Chakraborty C, Sharma AR, Bhattacharya M, et al. The Drug Repurposing for COVID-19 Clinical Trials Provide Very Effective Therapeutic Combinations: Lessons Learned From Major Clinical Studies. Front Pharmacol. 2021; 12:704205. doi:10.3389/fphar.2021.704205

22. Huang C, Wang Y, Li X, et al. Clinical features of patients infected with 2019 novel coronavirus in Wuhan, China. Lancet 2020, 395, 497–506.

23. Molla MA, Yeasmin M, Islam K, et al. Antibiotic Prescribing Patterns at COVID-19 Dedicated Wards in Bangladesh: Findings from a Single Center Study. Infect. PrevPract. 2021, 3, 100134.doi: 10.1016/j.infpip.2021.

24. Mah-E-Muneer S, Hassan MZ, Biswas Maaj, et al. Use of Antimicrobials among Suspected COVID-19 Patients at Selected Hospitals, Bangladesh: Findings from the First Wave of COVID-19 Pandemic. Antibiotics (Basel). 2021;10(6):738. doi: 10.3390/antibiotics10060738.

25. Knight GM, Glover RE, McQuaid CF, et al. Antimicrobial resistance and COVID-19: Intersections and implications. Elife. 2021;10:e64139. doi: 10.7554/eLife.64139.

26. Rawson TM, Moore LSP, Zhu N, et al. Bacterial and fungal co-infection inindividuals with coronavirus: A rapid review to supportCOVID-19 antimicrobial prescribing. Clin Infect Dis2020; 71(9):2459–2468.

27. Parveen M, Molla MA, Yeasmin M, et al. Evidences on Irrational Anti-Microbial Prescribing and Consumption among COVID-19 Positive Patients and Possible Mitigation Strategies: A Descriptive Cross Sectional Study. Bangladesh J. Infect. Dis. 2020, 7, S3–S7

28. Lind JN, Lovegrove MC, Geller AI, et al. Increase in Outpatient Ivermectin Dispensing in the US During the COVID-19 Pandemic: A Cross-Sectional Analysis. J Gen Intern Med. 202;18:1–3. doi: 10.1007/s11606-021-06948-6.

29. Nasir M, Chowdhury Asms, Zahan T. Self-medication during COVID-19 outbreak: A cross sectional online survey in Dhaka city. Int J Basic Clin Pharmacol 2020, 9:1325

30. Chen N, Zhou M, Dong X, et al. Epidemiological and clinical characteristics of 99 cases of 2019 novel coronavirus pneumonia in Wuhan, China: A descriptive study. Lancet 2020; 395: 507–513.

31. JoshiS, Parkar J, Ansari A, et al. Role of favipiravir in the treatment of COVID-19. Int J Infect Dis 2021;102:501–508

32. Raju R, Prajith V, Biatris PS, et al. Therapeutic role of corticosteroids in COVID-19: a systematic review of registered clinical trials. Futur J Pharm Sci 2021;7(1):67. doi:10.1186/s43094-021-00217-3

33. Wu R, Wang L, Kuo HD, et al. An Update on current therapeutic drugs treating COVID-19. Curr Pharmacol Rep. 2020;6(3):56–70.

34. Soni SL, Kajal K, Yaddanapudi LN, et al. Demographic & clinical profile of patients with COVID-19 at a tertiary care hospital in north India. Indian J Med Res 2021; 153 (1 & 2):115–125.

35. Barré J, Sabatier JM, Annweiler C. Montelukast Drug May Improve COVID-19 Prognosis: A Review of Evidence. Front Pharmacol. 2020;11: 1344. doi:10.3389/fphar.2020.01344

36. Brodlie M, Gupta A, Rodriguez-Martinez CE, et al. Leukotriene receptor antagonists as maintenance and intermittent therapy for episodic viral wheeze in children. Cochr DatabaseSyst Rev 2015, CD008202.

37. Mohan A, Tiwari, Bhatnagar S, et al. Clinico-demographic profile and hospital outcomes of COVID-19 patients admitted at atertiary care center in India. Indian J Med Res 2020; 152: 61–69.

38. Mondal RN, Sarker MAR, Das A, Ahsan MAK, Jahan SMS, Sultana A et al. Socio-demographic, clinical, hospital admission and oxygen requirement characteristics of COVID-19 patients of Bangladesh. 2020. medRxiv preprint doi: https://doi.org/10.1101/2020.08.14.20175018.t

39. Hossain HT, Chowdhury T, Majumder MI, et al. Demographic and clinical profile of 190 COVID-19 patients in a tertiary care private hospital of Dhaka, Bangladesh: an observational study. J Med 2020; 21: 82–8.

